# UROMODULIN AND RISK OF UPPER URINARY TRACT INFECTIONS: A MENDELIAN RANDOMIZATION STUDY

**DOI:** 10.1101/2024.06.18.24309082

**Authors:** Kristin Vardheim Liyanarachi, Helene Flatby, Stein Hallan, Bjørn Olav Åsvold, Jan Kristian Damås, Tormod Rogne

**Affiliations:** Mid-Norway Center for Sepsis Research, Department of Circulation and Medical Imaging, NTNU, Norwegian University of Science and Technology, Trondheim, Norway; Department of Infectious Diseases, Clinic of Medicine, St. Olav’s Hospital, Trondheim University Hospital, Trondheim, Norway; Department of Clinical and Molecular Medicine, NTNU, Norwegian University of Science and Technology, Trondheim, Norway; Department of Nephrology, Clinic of Medicine, St Olav’s Hospital, Trondheim University Hospital, Trondheim, Norway; HUNT Center for Molecular and Clinical Epidemiology, Department of Public Health and Nursing, NTNU, Norwegian University of Science and Technology, Trondheim, Norway; Department of Endocrinology, Clinic of Medicine, St. Olav’s Hospital, Trondheim University Hospital, Trondheim, Norway; HUNT Research Center, Department of Public Health and Nursing, NTNU, Norwegian University of Science and Technology, Levanger, Norway; Yale Department of Chronic Disease Epidemiology and Center for Perinatal, Pediatric and Environmental Epidemiology, Yale School of Public Health, New Haven, CT, USA

## Abstract

**Background:** Observational studies have suggested that uromodulin, produced by the kidneys, may reduce the risk of upper urinary tract infections, but are limited by potential confounding. To address this concern, we conducted a two-sample Mendelian randomization study to explore this association.

**Methods:** We identified uncorrelated (r^2^< 0.01) single nucleotide polymorphisms strongly associated (p<5 × 10^−6^) with urinary and serum uromodulin from two genome-wide association studies. Both studies accounted for kidney function. Genetic associations for the risk of upper urinary tract infections were extracted from an independent genome-wide association study. Inverse-variance weighted and sensitivity analyses were performed.

**Results:** The study included 29,315 and 13,956 participants with measured urinary and serum uromodulin, respectively, and 3,873 and 512,608 subjects with and without upper urinary tract infections. A one standard deviation increase in genetically predicted urinary uromodulin was associated with an odds ratio for upper urinary tract infections of 0.80 (95% confidence interval 0.67 to 0.95, p = 0.01). For serum uromodulin, a one standard deviation increase was associated with an odds ratio of 0.95 (95% confidence interval 0.89 to 1.01, p = 0.12). The results were consistent across sensitivity analyses.

**Conclusion:** In this two-sample mendelian randomization study we found that increased levels of genetically predicted urinary uromodulin were associated with a reduced risk of upper urinary tract infections. A similar trend was observed for serum uromodulin. Our findings align with results from traditional observational studies which together support that uromodulin may have a protective role against upper urinary tract infections

## INTRODUCTION

Upper urinary tract infection (UTI) is one of the most common causes of infection, is observed in all ages and sexes, and may develop into sepsis with high morbidity and mortality [1-3]. Thus, there is a need to identify targetable risk factors to reduce its disease burden and to ensure proper risk stratification.

Several known risk factors contribute to the susceptibility and severity of upper UTIs, including age, female sex, catheterization, and genetic predisposition [4-10]. In particular, chronic kidney failure increases the risk of upper UTIs. Recently, it has been suggested that this excess risk may be explained by the effects of uromodulin [11, 12].

Uromodulin (also referred to as Tamm-Horsfall protein) is a kidney-specific protein produced by the epithelial cells lining the ascending limb of the loop of Henle and in the distal convoluted tubule [11, 13]. Most of uromodulin is secreted into the urine, but it can also be measured in the systemic circulation [14]. Urine concentration is closely linked to a person’s number of nephrons and, hence, glomerular filtration rate [13]. The exact mechanisms of how uromodulin affects the risk for upper UTIs or other infections are unclear. However, uromodulin’s ability to block bacterial colonization of the urothelium is a possible explanation [15, 16]. In addition, uromodulin may be a regulator of NaCl transport processes and have an immunomodulatory role in the innate immune system by activating dendritic cells and other immunomodulatory cells [15, 17, 18]. Finally, genetic studies have found that variation in *UMOD*, the gene coding for uromodulin, is associated with a spectrum of rare and common kidney diseases [17, 19, 20], hypertension, and renal stones [16, 17, 19-21].

Observational studies have found that increasing levels of urinary uromodulin were associated with reduced risk of upper and lower UTIs [15, 17]. Conventional observational studies are, however, potentially limited by residual confounding (e.g., from comorbidities). Mendelian randomization (MR) has emerged as a technique that may limit such bias [22]. Because genetic variants are allocated randomly during gamete formation, they can be used as instruments (similar to the random allocation of a treatment in a trial), thus potentially limiting confounding [23].

In this two-sample MR study, we aimed to examine the relationship between genetically predicted urinary and serum uromodulin and the risk of upper UTIs.

## METHODS

### Study population

This MR study is reported according to Strengthening the Reporting of Observational Studies in Epidemiology Using Mendelian Randomization (STROBE-MR) guidelines [22] (Supplementary Table 1). The MR analyses were performed using individuals of primarily European ancestry. The summary-level data for the genetic associations for urinary uromodulin, serum uromodulin and upper UTIs were publicly available [20, 21, 24]. We used single nucleotide polymorphisms (SNPs) identified in these studies to assess how the genetic instruments of exposure (urinary and serum uromodulin) affected the risk of upper UTIs.

### Instrumental variable selection for uromodulin

Genetic instruments were extracted from the most comprehensive genome-wide association studies (GWASs) on urinary and serum uromodulin (Table 1) [20, 21]. The GWAS on urinary uromodulin was performed in adults from 13 different cohorts of European ancestry. The urinary uromodulin levels were indexed to urine creatinine, inverse-normal transformed, and adjusted for sex, age, and relatedness. The cohorts were representative of the general adult population, and the participants delivered urinary samples upon inclusion, in addition to blood samples for creatinine and genotyping.

**Table 1.** Genome-Wide Association Studies Used as Sources for Two-Sample Mendelian Randomization Analyses

|  | Trait | Ancestry | Countries | Cohorts used | Phenotype definition | Cases/controls or number of participants | No. of SNPs | % of variance explained |
| --- | --- | --- | --- | --- | --- | --- | --- | --- |
| **Joseph et al<sup>[20]</sup>** | urinary uromodulin | European | Canada, Switzerland, Croatia, USA, Liechtenstein, Germany, United Kingdom, Italy, | CARTaGENE, CoLaus, CROATIA-Korcula, CROATIA-Split, CROATIA-Vis, Framingham Heart Study, Genetic and Phenotypic Determinants of Blood Pressure and other Cardiovascular Risk Factors, German Chronic Kidney Disease, Generation Scotland: Scottish Family Health Study, INGI-Varlantino, INGI-Val Borbera, Lothian Birth Cohort 1936, Viking Health Study-Shetland | Unit of exposure was per standard deviation increase of urinary uromodulin indexed to creatinine and adjusted for sex, age, and relatedness. | 29,315 | 17 | 3.1 |
| **Li et al<sup>[21]</sup>** | serum uromodulin | 81.1% European, 2.9% African American, 15.9% Hispanic | USA, Germany, Canada | Cardiovascular Health Study, German Chronic Kidney Disease, Cooperative Health Research in the Region Augsburg, Ludwigshafen Risk and Cardiovascular, Outcome Reduction with an Initial Glargine Intervention | Unit of exposure was per standard deviation increase of serum uromodulin adjusted for eGFR, age and sex | 13,956 | 24 | 24.3 |
| **Flatby et al<sup>[24]</sup>** | Upper UTI | European | United Kingdom, Norway, USA | UK Biobank, The Trøndelag Health Study, Michigan Genomics Initiative | **Cases:** ICD-9 code 590 (infections of the kidney), ICD-10 codes N10 (acute pyelonephritis) and N12 (tubulo-interstitial nephritis, not specified as acute or chronic). **Controls:** Subjects not admitted with the above codes. Analysis adjusted for age, sex and ancestry | **Both sexes;** 3,873/512,608<br>**Women:** 1,831/255,541<br>**Men:** 896/219,040 | 8<sup>a</sup> | N/A |

a) only used in the bidirectional analysis
Abbreviations: No; number, SNP; single nucleotide polymorphism, ICD; International classification of disease, N/A; not applicable

The GWAS on serum uromodulin included five prospective studies [21]. Two of them had a population-based design, while the other three included patients with chronic kidney disease, cardiovascular disease, and impaired glucose tolerance/early type 2 diabetes respectively. The majority (11,314 [81.1%]) of the participants were of European ancestry, while a small proportion where of Hispanic and African American ancestry (2,216 [15.9%] and 400 [2.9%], respectively). The unit used in the analysis was rank-based inverse normal transformed residuals of uromodulin, and it was adjusted for estimated glomerular filtration rate (eGFR), age, and sex.

For both urinary (main analysis) and serum (sub analysis) uromodulin, we extracted SNPs that were strongly associated with the exposure, defined as *p* ≤ 5 × 10^−6^. This threshold is less stringent than what is often used in Mendelian randomization studies (*p* ≤ 5 × 10^−8^) [25], but it was applied to identify a sufficient number of genetic instruments. In a sensitivity analysis of urinary uromodulin, we also performed the MR analyses using the *p* ≤ 5 × 10^−8^ threshold. To ensure that the included SNPs were independent of one another, we assessed 10,000 kb windows and when the *r*^2^ was 0.01 between two SNPs, we kept the SNP with the lowest p-value; the 1,000 Genomes Project on European ancestry was used as reference population [26].

### Genetic associations of liability to upper urinary tract infections

Genetic associations of liability to upper UTIs were extracted from a GWAS using data from the UK Biobank, The Trøndelag Health Study (HUNT), and the Michigan Genomic Initiative [24]. Cases were defined based on hospital discharge codes (Table 1), and the study included 3,873 cases and 512,608 controls. There is no known overlap of participants between the outcome and exposure GWASs, however it is possible that some of the participants in the Scottish Family Health study could also have joined the UK Biobank, as these two collected patient data within the same time-span and geographical area.

### Statistical analyses

The strength of each genetic instrument was estimated using the F statistic: F = R^2^(N − 2)/(1 − R^2^), where R^2^ equals the proportion of variance explained by the genetic instrument, and N is the effective sample size of the GWAS for the SNP-exposure association. The R^2^ value was calculated using the formula 2 x MAF(1 − MAF)beta^2^, where beta represents the effect estimate of the genetic variant in the exposure, and MAF represents the minor allele frequency [27] (Supplementary Table 2).

Before performing the analyses, the exposure and outcome files were harmonized to ensure that all datasets evaluated the effect of the same allele for the same SNP. We then calculated the Wald ratio for each SNP, defined as the association between the SNP and upper UTIs divided by the association between the SNP and uromodulin. The Wald ratios for all SNPs were next summarized using inverse-variance weighting (IVW) which provides an unbiased estimate if the instruments are valid.

An instrument is valid if it is associated with the risk factor under study, shares no common cause with the outcome, and only affects the outcome through the risk factor [23]. To examine the validity of the MR analyses, we conducted a wide range of sensitivity analyses: The weighted median, weighted mode, simple mode, MR Egger regression, and the MR Egger intercept test. These sensitivity analyses estimate the associations under different assumptions about horizontal pleiotropy, which occurs when genetic variants are associated with multiple phenotypes through an independent pathway other than the exposure [28]. A consistent result across these sensitivity and IVW analysis supports that the instrumental variable assumptions are not violated. Another set of sensitivity analyses included the Cochran’s Q statistical test to evaluate the presence of heterogeneity between the Wald ratios [29] and leave-one-out analyses to examine whether the overall finding was driven by single SNPs [30].

We further evaluated whether the instruments were associated with other phenotypes that might introduce bias (e.g., horizontal pleiotropy) using the Open Targets Genetics platform [31].

Since sex-combined MR analysis could potentially mask different effects by sex, either in direction or magnitude, and because we know that there are etiological differences between men and women for the risk of upper UTIs, we also conducted sex-stratified analyses (Table 1).

Lastly, to assess the possibility of reverse causation, we conducted a bidirectional analysis, where we used eight independent SNPs strongly associated with upper UTIs as exposure (same selection criteria as for the uromodulin instruments) [24]. The genetic association of urinary uromodulin from the GWAS was used in the main analysis as the outcome [20].

All analyses were conducted using the TwoSample MR package version 0.5.7. in R version 4.1.2.

### Ethics

We only used summary-level data from studies with relevant participant consent and ethical approval, and ethical approval from an institutional review board was therefore not necessary for the present study.

## RESULTS

In the main analysis we identified 17 SNPs as genetic instruments for urinary uromodulin, explaining 3.1% of its variance, while 24 SNPs explained 24.3% of the variance of serum uromodulin in the sub analysis. In the sensitivity analysis using the more stringent threshold of *p* ≤ 5×10^−8^, we identified two SNPs for urinary uromodulin explaining 1.9% of the variance (Supplementary Table 2). The F-statistic was > 10 for all genetic instruments.

In the IVW analysis, a one standard deviation increase in genetically predicted urinary uromodulin was associated with an odds ratio for upper UTI of 0.80 (95% CI 0.67 to 0.95, p = 0.01). (Figure 1 and Supplementary Figure 1). Similar results were observed when we used the stricter statistical p-value threshold (Figure 1).

**Figure 1.**
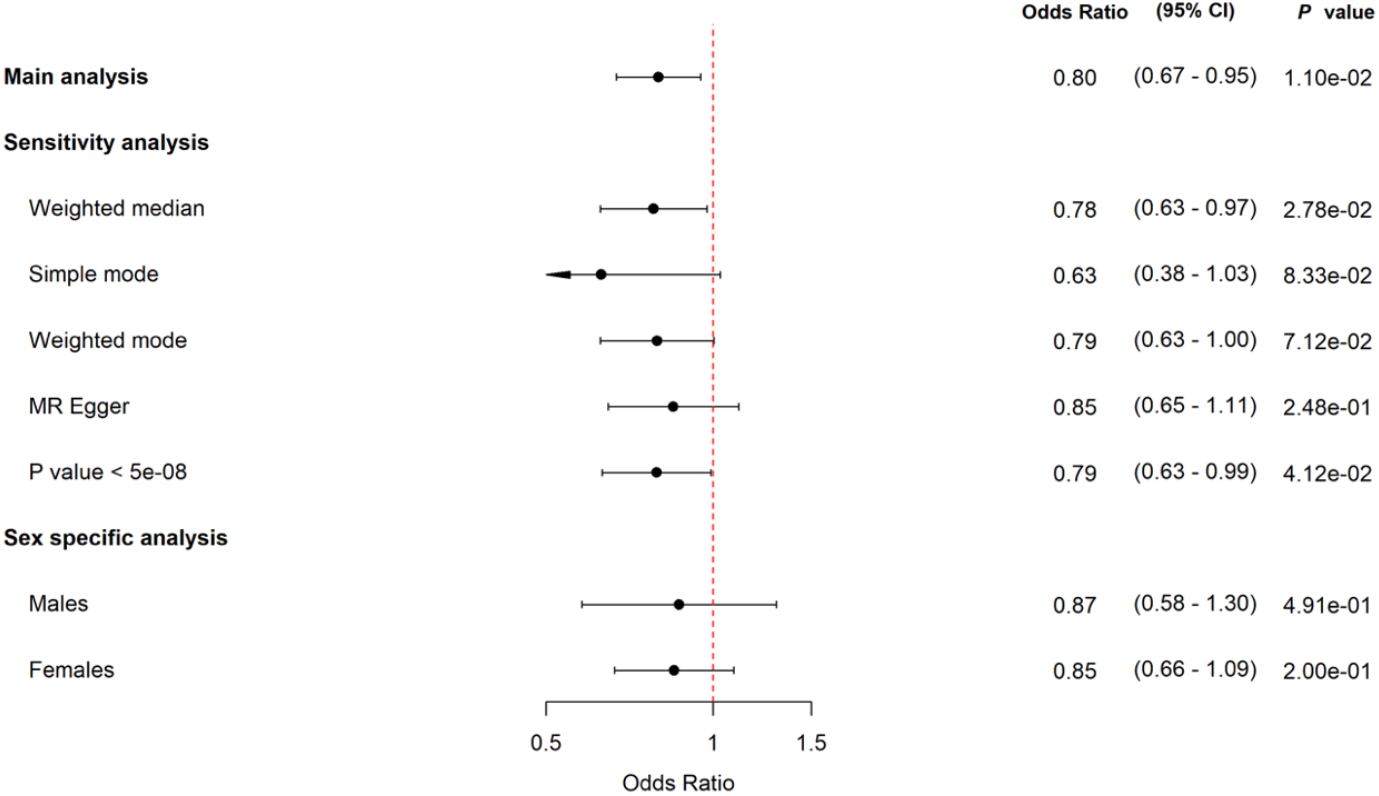
Mendelian randomization analyses of urinary uromodulin and risk of upper UTIs. Forest plot of the main analysis and the sensitivity and sex-specific analyses of the association between genetically predicted urinary uromodulin and risk of upper UTI. The horizontal bars represent 95% CI. Abbreviations: CI; confidence intervals.

There was a tendency for a negative association between serum uromodulin and risk of upper UTIs, where a one standard deviation increase in genetically predicted serum uromodulin was associated with an odds ratio for upper UTI of 0.95 (95% CI 0.89 to 1.01, p = 0.12) (Supplementary Figure 2).

The MR-Egger, weighted median, and mode-based sensitivity analyses supported the findings from the IVW analysis (Figure 1), and MR Egger intercept test did not suggest bias due to pleiotropy (p = 0.66). No heterogeneity between the genetic variants was observed (Cochran’s Q test p = 0.87). In the leave-one-out analysis, the observed association of genetically predicted urinary uromodulin with upper UTIs did not change meaningfully, even when the strongest genetic instrument located in the *UMOD* gene was omitted (Supplementary Table 3).

In the bidirectional MR analysis, there was no association between genetic liability to upper UTIs and levels of urinary uromodulin (OR 1.02, 95% CI 0.94 to 1.10, p=0.67)

Apart from the SNP in the *UMOD* gene, which is important for different renal diseases, none of the other SNPs used as genetic instruments were associated with traits that are plausible to bias the association between uromodulin and risk of upper UTIs.

Finally, in the sex-stratified analyses, we found similar results (OR 0.87, 95% CI 0.58 to 1.30, p=0.49 in men and OR 0.85, 95% CI 0.66 to 1.09, p= 0.20 in women (Figure 1).

## DISCUSSION

In this two-sample MR study, we found that higher genetically predicted urinary uromodulin concentrations were associated with lower risk of upper UTIs. This finding was robust across a range of sensitivity analyses.

Our findings add to the increasing body of literature reporting an association between uromodulin and risk of UTIs. A previous study showed an inverse correlation between urinary uromodulin levels and local and systemic markers of UTIs (upper and lower UTIs combined), concluding that urinary uromodulin could have a protective effect in the general population [17]. The authors also discuss that the SNPs identified in the *UMOD* gene – associated with a higher urinary excretion of uromodulin – may have been kept at high frequency in the human population due to its protective effect against UTIs and subsequently reproductive function. A prospective study of predominantly outpatients also observed a negative correlation between urinary uromodulin and lower UTIs [15].

A phenome-wide association study explored associations between the most important SNPs predicting higher levels of urinary uromodulin and 1,528 different clinical diagnosis codes [32]. They concluded that these genetic variants were associated with lower odds of UTIs (both upper and lower) and urinary tract calculus. Our results further add to these findings, by including a larger set of SNPs, evaluating potential biases due to pleiotropic effects, and by considering upper UTIs independently.

The search for relevant biomarkers in severe infections is ongoing, and our study suggests that the role of uromodulin in predicting upper UTIs should be examined further. Further, understanding of mechanisms that increase uromodulin concentrations or prevent degradation of uromodulin could potentially guide the development of new treatment options for upper UTIs [15]. For example, a new class of small-molecular-weight compounds known as mannosides is being developed to treat and prevent UTIs [33, 34]. These agents inhibit bacterial colonization in the uroepithelium using a mechanism very similar to uromodulin [18]. The role as a biomarker would be of further importance if the level of circulating serum uromodulin was found to play a role. While there was a tendency for a protective association of increasing serum uromodulin and risk of upper UTIs, this was not significant. Earlier studies have shown that serum uromodulin protects against sepsis mortality in mice [14]. A recently published MR study found no association between genetically determined serum uromodulin concentration and sepsis or severe pneumonia [16]. Further studies are needed to evaluate whether measuring urinary or serum uromodulin levels can be of prognostic value when assessing patients with upper UTIs.

This study has several strengths and limitations. By choosing an MR design, we greatly reduced the risk of confounding, which often affects observational studies. Furthermore, the sensitivity analyses did not indicate any bias due to pleiotropy. However, bias due to pleiotropic effects is not possible to completely rule out. In our study, this could be the case with chronic kidney disease, leading to reduced nephron mass and, hence a lower level of urinary uromodulin [13]. The urinary uromodulin levels used in the GWAS were indexed to creatinine and the serum uromodulin levels were adjusted for eGFR. This should address most of the potential pleiotropic effects through kidney function, although there may be some bias from kidney function not captured by creatinine or eGFR. The sensitivity and subgroup analyses were affected by low statistical power. The analyses were performed on participants predominantly of European ancestry, decreasing the generalizability of our findings to the world’s other populations. A benefit of using subjects of the same ancestry group is that it reduces the risk of confounding due to population characteristics. Also, the exposure and outcome data used were from separate populations, greatly reducing the risk of confounding bias due to overlapping samples.

## CONCLUSIONS

Our results of an association between higher genetically predicted urinary uromodulin levels and lower risk of upper UTIs support previous traditional observational studies’ findings. This strengthens the likelihood that urinary uromodulin may play a biological role in the susceptibility to upper UTIs. Further research into potential therapeutic opportunities and the possible utility of urinary uromodulin as a diagnostic marker is warranted.

## Data Availability

All data produced in the present study are available upon reasonable request to the authors

## FUNDING

This work was supported by Samarbeidsorganet Helse Midt-Norge, NTNU (Norwegian University of Science and Technology) (Trondheim, Norway) (KL). Open access funding provided by NTNU Norwegian University of Science and Technology (incl St. Olavs Hospital - Trondheim University Hospital). TR was funded by CTSA Grant Number UL1 TR001863 from the National Center for Advancing Translational Science (NCATS), a component of the National Institutes of Health (NIH). The contents of this manuscript are solely the responsibility of the authors and do not necessarily represent the official views of NIH. None of the authors or their institutions have at any time received payment or services from a third party for any aspect of the submitted work.

## ACKNOWLEDGEMENTS

We want to acknowledge the participants and investigators of alle the included published GWASs.

## CONFLICTS OF INTEREST

Kristin Vardheim Liyanarachi: no conflict, Helene Flatby: no conflict, Stein Hallan: no conflict, Bjørn O Åsvold: no conflict, Jan Kristian Damås: no conflict, Tormod Rogne: no conflict. the authors or their institutions have not received any payments or services in the past 36 months from a third party that could be perceived to influence, or give the appearance of potentially influencing, the submitted work.

## Notes

### Competing Interest Statement

The authors have declared no competing interest.

### Author Declarations

We only used summary-level data from studies with relevant participant consent and ethical approval, and ethical approval from an institutional review board was therefore not necessary for the present study. Links to published GWASs: doi: 10.1681/ASN.2021040491.doi: 10.1172/jci.insight.157035 doi: 10.1093/infdis/jiae231

## REFERENCES

1. Foxman B. The epidemiology of urinary tract infection. Nature Reviews Urology 2010; 7:653–60.

2. Rudd KE, Johnson SC, Agesa KM, et al. Global, regional, and national sepsis incidence and mortality, 1990-2017: analysis for the Global Burden of Disease Study. Lancet 2020; 395:200–11.

3. Zeng Z, Zhan J, Zhang K, Chen H, Cheng S. Global, regional, and national burden of urinary tract infections from 1990 to 2019: an analysis of the global burden of disease study 2019. World J Urol 2022; 40:755–63.

4. Kaur R, Kaur R. Symptoms, risk factors, diagnosis and treatment of urinary tract infections. Postgraduate Medical Journal 2020; 97:803–12.

5. Salvatore S, Salvatore S, Cattoni E, et al. Urinary tract infections in women. Eur J Obstet Gynecol Reprod Biol 2011; 156:131–6.

6. Dielubanza EJ, Schaeffer AJ. Urinary tract infections in women. Med Clin North Am 2011; 95:27–41.

7. Liyanarachi KV, Solligård E, Mohus RM, Åsvold BO, Rogne T, Damås JK. Incidence, recurring admissions and mortality of severe bacterial infections and sepsis over a 22-year period in the population-based HUNT study. PLOS ONE 2022; 17:e0271263.

8. Nicolle LE. Catheter associated urinary tract infections. Antimicrob Resist Infect Control 2014; 3:23.

9. Scholes D, Hawn TR, Roberts PL, et al. Family history and risk of recurrent cystitis and pyelonephritis in women. J Urol 2010; 184:564–9.

10. Zaffanello M, Malerba G, Cataldi L, et al. Genetic risk for recurrent urinary tract infections in humans: a systematic review. J Biomed Biotechnol 2010; 2010:321082.

11. Weiss GL, Stanisich JJ, Sauer MM, et al. Architecture and function of human uromodulin filaments in urinary tract infections. Science 2020; 369:1005–10.

12. Foley RN. Infections in patients with chronic kidney disease. Infect Dis Clin North Am 2007; 21:659-72, viii.

13. Devuyst O, Pattaro C. The UMOD Locus: Insights into the Pathogenesis and Prognosis of Kidney Disease. Journal of the American Society of Nephrology 2018; 29:713–26.

14. LaFavers KA, Hage CA, Gaur V, et al. The kidney protects against sepsis by producing systemic uromodulin. Am J Physiol Renal Physiol 2022; 323:F212–f26.

15. Garimella PS, Bartz TM, Ix JH, et al. Urinary Uromodulin and Risk of Urinary Tract Infections: The Cardiovascular Health Study. Am J Kidney Dis 2017; 69:744–51.

16. Eriksson M, Lipcsey M, Ilboudo Y, Yoshiji S, Richards B, Hultström M. Uromodulin in sepsis and severe pneumonia - A two-sample Mendelian randomization study. Physiol Genomics 2024.

17. Ghirotto S, Tassi F, Barbujani G, et al. The Uromodulin Gene Locus Shows Evidence of Pathogen Adaptation through Human Evolution. J Am Soc Nephrol 2016; 27:2983–96.

18. Säemann MD, Hörl WH, Weichhart T. Uncovering host defences in the urinary tract: cathelicidin and beyond. Nephrol Dial Transplant 2007; 22:347–9.

19. Schaeffer C, Devuyst O, Rampoldi L. Uromodulin: Roles in Health and Disease. Annu Rev Physiol 2021; 83:477–501.

20. Joseph CB, Mariniello M, Yoshifuji A, et al. Meta-GWAS Reveals Novel Genetic Variants Associated with Urinary Excretion of Uromodulin. J Am Soc Nephrol 2022; 33:511–29.

21. Li Y, Cheng Y, Consolato F, et al. Genome-wide studies reveal factors associated with circulating uromodulin and its relationships to complex diseases. JCI Insight 2022; 7.

22. Skrivankova VW, Richmond RC, Woolf BAR, et al. Strengthening the Reporting of Observational Studies in Epidemiology Using Mendelian Randomization: The STROBE-MR Statement. JAMA 2021; 326:1614–21.

23. Davies NM, Holmes MV, Davey Smith G. Reading Mendelian randomisation studies: a guide, glossary, and checklist for clinicians. Bmj 2018; 362:k601.

24. Flatby HM, Ravi A, Liyanarachi KV, et al. A genome-wide association study of susceptibility to upper urinary tract infections. J Infect Dis 2024.

25. Burgess S, Davey Smith G, Davies N, et al. Guidelines for performing Mendelian randomization investigations: update for summer 2023 [version 3; peer review: 2 approved]. Wellcome Open Research 2023; 4.

26. Auton A, Brooks LD, Durbin RM, et al. A global reference for human genetic variation. Nature 2015; 526:68–74.

27. Burgess S, Dudbridge F, Thompson SG. Combining information on multiple instrumental variables in Mendelian randomization: comparison of allele score and summarized data methods. Stat Med 2016; 35:1880–906.

28. Sanderson E, Glymour MM, Holmes MV, et al. Mendelian randomization. Nature Reviews Methods Primers 2022; 2:1–21.

29. Bowden J, Davey Smith G, Burgess S. Mendelian randomization with invalid instruments: effect estimation and bias detection through Egger regression. Int J Epidemiol 2015; 44:512–25.

30. Hemani G, Bowden J, Davey Smith G. Evaluating the potential role of pleiotropy in Mendelian randomization studies. Hum Mol Genet 2018; 27:R195–R208.

31. Ghoussaini M, Mountjoy E, Carmona M, et al. Open Targets Genetics: systematic identification of trait-associated genes using large-scale genetics and functional genomics. Nucleic Acids Research 2020; 49:D1311–D20.

32. Akwo EA, Chen H-C, Liu G, et al. Phenome-Wide Association Study of UMOD Gene Variants and Differential Associations With Clinical Outcomes Across Populations in the Million Veteran Program a Multiethnic Biobank. Kidney International Reports 2022; 7:1802–18.

33. Cusumano CK, Pinkner JS, Han Z, et al. Treatment and prevention of urinary tract infection with orally active FimH inhibitors. Science translational medicine 2011; 3:109ra15–ra15.

34. Wellens A, Garofalo C, Nguyen H, et al. Intervening with Urinary Tract Infections Using Anti-Adhesives Based on the Crystal Structure of the FimH–Oligomannose-3 Complex. PLOS ONE 2008; 3:e2040.

