## Supplementary Figure 1 for "UROMODULIN AND RISK OF UPPER URINARY TRACT INFECTIONS: A MENDELIAN RANDOMIZATION STUDY"

**Supplementary Figure 1:** Scatter plot of MR analysis on urinary uromodulin and upper UTI

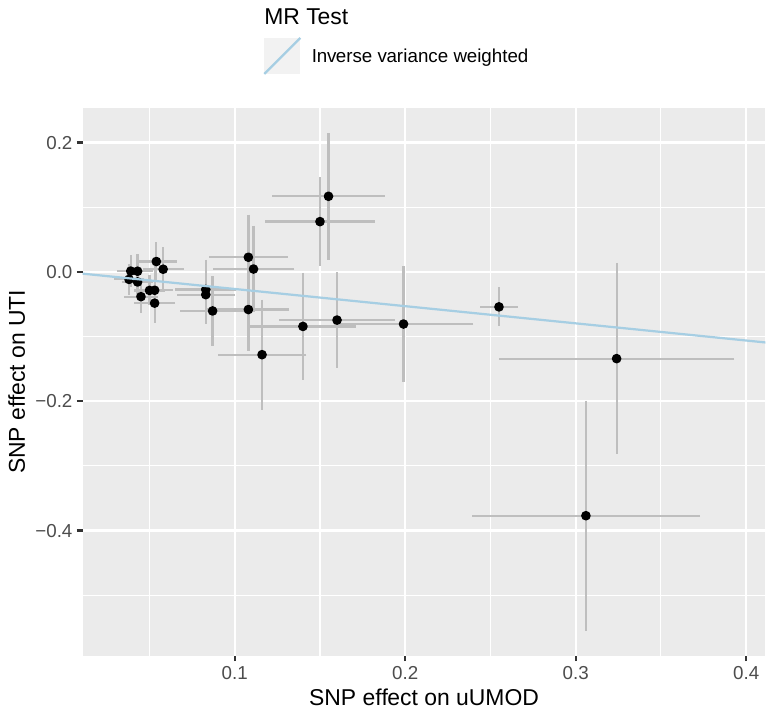

*Legend*: Scatter plot of the MR analysis (IVW) (*r^2^* < 0.01 within 10,000 kb windows and *P* ≤ 5E-06) using urinary uromodulin as exposure and upper UTI as outcome.
