## Supplementary Figure 2 for "UROMODULIN AND RISK OF UPPER URINARY TRACT INFECTIONS: A MENDELIAN RANDOMIZATION STUDY"

**Supplementary Figure 2:** Scatter plot of MR analysis on serum uromodulin and upper UTI

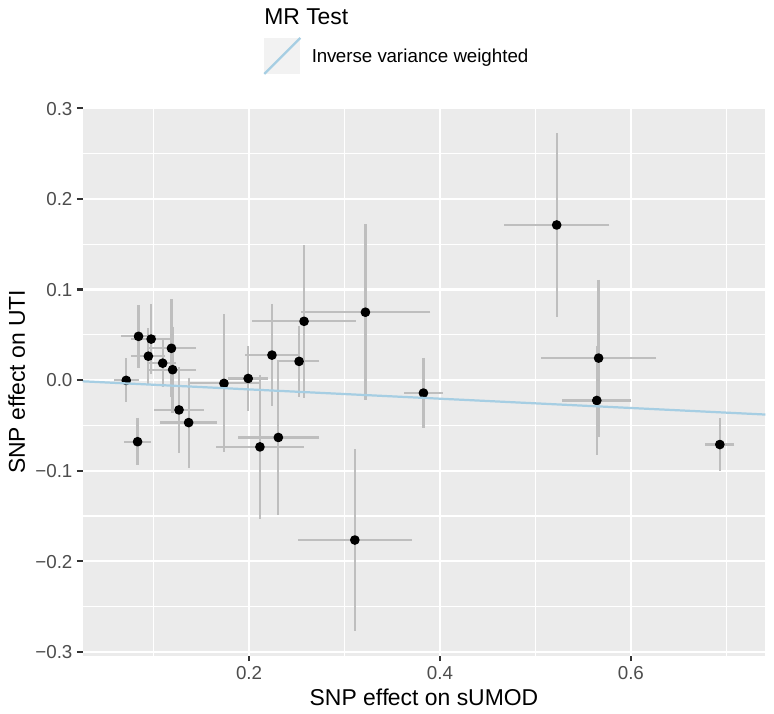

*Legend*: Scatter plot of the MR analysis (IVW) (*r^2^* < 0.01 within 10,000 kb windows and *P* ≤ 5E-06) using serum uromodulin as exposure and upper urinary UTI as outcome.
