## Supplementary Table 1 for "UROMODULIN AND RISK OF UPPER URINARY TRACT INFECTIONS: A MENDELIAN RANDOMIZATION STUDY"

**Supplementary Table 1:** STROBE-MR Checklist of Recommended Items to Address in Reports of Mendelian Randomization Studies.

| **Item No.** | **Section** | **Checklist item** | **Page No.** | **Relevant text from manuscript** |
| --- | --- | --- | --- | --- |
| 1 | **TITLE and ABSTRACT** | Indicate Mendelian randomization (MR) as the study’s design in the title and/or the abstract if that is a main purpose of the study | 1 | Uromodulin and risk of upper urinary tract infections: A Mendelian randomization study. |
|  | **INTRODUCTION** |  |  |  |
| 2 | **Background** | Explain the scientific background and rationale for the reported study. What is the exposure? Is a potential causal relationship between exposure and outcome plausible? Justify why MR is a helpful method to address the study question | 3 | Observational studies have found that increasing levels of urinary uromodulin were associated with reduced risk of upper and lower UTIs. Conventional observational studies are, however, potentially limited by residual confounding (e.g., from comorbidities). Mendelian randomization (MR) has emerged as a technique that may limit such bias. Because genetic variants are allocated randomly during gamete formation, they can be used as instruments (similar to the random allocation of a treatment in a trial), thus potentially limiting confounding.  In this two-sample MR study, we aimed to examine the relationship between genetically predicted urinary and serum uromodulin and the risk of upper UTIs. |
| 3 | **Objectives** | State specific objectives clearly, including pre-specified causal hypotheses (if any). State that MR is a method that, under specific assumptions, intends to estimate causal effects | 3+6 | In this two-sample MR study, we aimed to examine the relationship between genetically predicted urinary and serum uromodulin and the risk of upper UTIs. Observational studies have found that increasing levels of urinary uromodulin were associated with reduced risk of upper and lower UTIs. Conventional observational studies are, however, potentially limited by residual confounding (e.g., from comorbidities). Mendelian randomization (MR) has emerged as a technique that may limit such bias. An instrument is valid if it is associated with the risk factor under study, shares no common cause with the outcome, and only affects the outcome through the risk factor. |
|  | **METHODS** |  |  |  |
| 4 | **Study design and data sources** | Present key elements of the study design early in the article. Consider including a table listing sources of data for all phases of the study. For each data source contributing to the analysis, describe the following: | 4 | Table 1 Genome-Wide Association Studies Used as Sources for two-Sample Mendelian Randomization Analyses |
|  | a) | Setting: Describe the study design and the underlying population, if possible. Describe the setting, locations, and relevant dates, including periods of recruitment, exposure, follow-up, and data collection, when available. | 4-6 | Genetic instruments were extracted from the most comprehensive genome-wide association studies (GWASs) on urinary and serum uromodulin. The GWAS on urinary uromodulin was performed in adults from 13 different cohorts of European ancestry. The urinary uromodulin levels were indexed to urine creatinine, inverse-normal transformed, and adjusted for sex, age, and relatedness. The cohorts were representative of the general adult population, and the participants delivered urinary samples upon inclusion, in addition to blood samples for creatinine and genotyping.  Genetic associations of liability to upper UTIs were extracted from a GWAS using data from the UK Biobank, The Trøndelag Health Study (HUNT), and the Michigan Genomic Initiative. Cases were defined based on hospital discharge codes, and the study included 3,873 cases and 512,608 controls. There is no known overlap of participants between the outcome and exposure GWASs, however it is possible that some of the participants in the Scottish Family Health study could also have joined the UK Biobank, as these two collected patient data within the same time-span and geographical area.  . |
|  | b) | Participants: Give the eligibility criteria, and the sources and methods of selection of participants. Report the sample size, and whether any power or sample size calculations were carried out prior to the main analysis | 4-6 | See above |
|  | c) | Describe measurement, quality control and selection of genetic variants | 4-5 | Genetic instruments were extracted from the most comprehensive genome-wide association studies (GWASs) on urinary and serum uromodulin. The GWAS on urinary uromodulin was performed in adults from 13 different cohorts of European ancestry. The urinary uromodulin levels were indexed to urine creatinine, inverse-normal transformed, and adjusted for sex, age, and relatedness. The cohorts were representative of the general adult population, and the participants delivered urinary samples upon inclusion, in addition to blood samples for creatinine and genotyping.  Genetic associations of liability to upper UTIs were extracted from a GWAS using data from the UK Biobank, The Trøndelag Health Study (HUNT), and the Michigan Genomic Initiative Cases were defined based on hospital discharge codes, and the study included 3,873 cases and 512,608 controls. There is no known overlap of participants between the outcome and exposure GWASs, however it is possible that some of the participants in the Scottish Family Health study could also have joined the UK Biobank, as these two collected patient data within the same time-span and geographical area. |
|  | d) | For each exposure, outcome, and other relevant variables, describe methods of assessment and diagnostic criteria for diseases | 5 | Table 1 |
|  | e) | Provide details of ethics committee approval and participant informed consent, if relevant | 7 | We only used summary-level data from studies with relevant participant consent and ethical approval, and ethical approval from an institutional review board was therefore not necessary for the present study. |
| 5 | **Assumptions** | Explicitly state the three core IV assumptions for the main analysis (relevance, independence and exclusion restriction) as well assumptions for any additional or sensitivity analysis | 7 | The strength of each genetic instrument was estimated using the F statistic: F = R^2^(N − 2)/(1 − R^2^), where R^2^ equals the proportion of variance explained by the genetic instrument, and N is the effective sample size of the GWAS for the SNP-exposure association. The R^2^ value was calculated using the formula 2 x MAF(1 − MAF)beta^2^, where beta represents the effect estimate of the genetic variant in the exposure, and MAF represents the minor allele frequency. An instrument is valid if it is associated with the risk factor under study, shares no common cause with the outcome, and only affects the outcome through the risk factor. To examine the validity of the MR analyses, we conducted a wide range of sensitivity analyses: The weighted median, weighted mode, simple mode, MR Egger regression, and the MR Egger intercept test. These sensitivity analyses estimate the associations under different assumptions about horizontal pleiotropy, which occurs when genetic variants are associated with multiple phenotypes through an independent pathway other than the exposure. A consistent result across these sensitivity and IVW analysis supports that the instrumental variable assumptions are not violated. Another set of sensitivity analyses included the Cochran's Q statistical test to evaluate the presence of heterogeneity between the Wald ratios and leave-one-out analyses to examine whether the overall finding was driven by single SNPs. |
| 6 | **Statistical methods: main analysis** | Describe statistical methods and statistics used |  |  |
|  | a) | Describe how quantitative variables were handled in the analyses (i.e., scale, units, model) | 4 | The urinary uromodulin levels were indexed to urine creatinine, inverse-normal transformed, and adjusted for sex, age, and relatedness. The unit used in the analysis was rank-based inverse normal transformed residuals of uromodulin, and it was adjusted for estimated glomerular filtration rate (eGFR), age, and sex.  The s-uromodulin levels were expressed as ng/ml. |
|  | b) | Describe how genetic variants were handled in the analyses and, if applicable, how their weights were selected | NA |  |
|  | c) | Describe the MR estimator (e.g. two-stage least squares, Wald ratio) and related statistics. Detail the included covariates and, in case of two-sample MR, whether the same covariate set was used for adjustment in the two samples | 6 | Before performing the analyses, the exposure and outcome files were harmonized to ensure that all datasets evaluated the effect of the same allele for the same SNP. We then calculated the Wald ratio for each SNP, defined as the association between the SNP and upper UTIs divided by the association between the SNP and uromodulin. The Wald ratios for all SNPs were next summarized using inverse-variance weighting (IVW) which provides an unbiased estimate if the instruments are valid.  Adjustments described in Table 1 |
|  | d) | Explain how missing data were addressed | NA |  |
|  | e) | If applicable, indicate how multiple testing was addressed | NA |  |
| 7 | **Assessment of assumptions** | Describe any methods or prior knowledge used to assess the assumptions or justify their validity | 6 | The strength of each genetic instrument was estimated using the F statistic: F = R2(N − 2)/(1 − R2), where R2 equals the proportion of variance explained by the genetic instrument, and N is the effective sample size of the GWAS for the SNP-exposure association. The R2 value was calculated using the formula 2 x MAF(1 − MAF)beta2, where beta represents the effect estimate of the genetic variant in the exposure, and MAF represents the minor allele frequency.  An instrument is valid if it is associated with the risk factor under study, shares no common cause with the outcome, and only affects the outcome through the risk factor. To examine the validity of the MR analyses, we conducted a wide range of sensitivity analyses: The weighted median, weighted mode, simple mode, MR Egger regression, and the MR Egger intercept test. These sensitivity analyses estimate the associations under different assumptions about horizontal pleiotropy, which occurs when genetic variants are associated with multiple phenotypes through an independent pathway other than the exposure. A consistent result across these sensitivity and IVW analysis supports that the instrumental variable assumptions are not violated. Another set of sensitivity analyses included the Cochran's Q statistical test to evaluate the presence of heterogeneity between the Wald ratios and leave-one-out analyses to examine whether the overall finding was driven by single SNPs. |
| 8 | **Sensitivity analyses and additional analyses** | Describe any sensitivity analyses or additional analyses performed (e.g. comparison of effect estimates from different approaches, independent replication, bias analytic techniques, validation of instruments, simulations) | 7 | An instrument is valid if it is associated with the risk factor under study, shares no common cause with the outcome, and only affects the outcome through the risk factor. To examine the validity of the MR analyses, we conducted a wide range of sensitivity analyses: The weighted median, weighted mode, simple mode, MR Egger regression, and the MR Egger intercept test. These sensitivity analyses estimate the associations under different assumptions about horizontal pleiotropy, which occurs when genetic variants are associated with multiple phenotypes through an independent pathway other than the exposure. A consistent result across these sensitivity and IVW analysis supports that the instrumental variable assumptions are not violated. Another set of sensitivity analyses included the Cochran's Q statistical test to evaluate the presence of heterogeneity between the Wald ratios and leave-one-out analyses to examine whether the overall finding was driven by single SNPs.  We further evaluated whether the instruments were associated with other phenotypes that might introduce bias (e.g., horizontal pleiotropy) using the Open Targets Genetics platform. |
| 9 | **Software and pre-registration** |  |  |  |
|  | a) | Name statistical software and package(s), including version and settings used | 8 | All MR analyses were conducted using the TwoSample MR package version 0.5.7. in R version 4.1.2. |
|  | b) | State whether the study protocol and details were pre-registered (as well as when and where) |  | No. |
|  | **RESULTS** |  |  |  |
| 10 | **Descriptive data** |  |  |  |
|  | a) | Report the numbers of individuals at each stage of included studies and reasons for exclusion. Consider use of a flow diagram |  | Supplementary table 2 |
|  | b) | Report summary statistics for phenotypic exposure(s), outcome(s), and other relevant variables (e.g. means, SDs, proportions) |  | Supplementarty table 2 |
|  | c) | If the data sources include meta-analyses of previous studies, provide the assessments of heterogeneity across these studies | NA |  |
|  | d) | For two-sample MR:  i.  Provide justification of the similarity of the genetic variant-exposure associations between the exposure and outcome samples  ii.  Provide information on the number of individuals who overlap between the exposure and outcome studies | 6 | All European ancestry  ii. There is no known overlap of participants between the outcome and exposure GWASs, however it is possible that some of the participants in the Scottish Family Health study could also have joined UK Biobank, as these two collected patient data within the same time-span and geographical area.  . |
| 11 | **Main results** |  |  |  |
|  | a) | Report the associations between genetic variant and exposure, and between genetic variant and outcome, preferably on an interpretable scale |  | Supplementary table 2 |
|  | b) | Report MR estimates of the relationship between exposure and outcome, and the measures of uncertainty from the MR analysis, on an interpretable scale, such as odds ratio or relative risk per SD difference |  | Figure 1 |
|  | c) | If relevant, consider translating estimates of relative risk into absolute risk for a meaningful time period | NA |  |
|  | d) | Consider plots to visualize results (e.g. forest plot, scatterplot of associations between genetic variants and outcome versus between genetic variants and exposure) |  | Figure 1. |
| 12 | **Assessment of assumptions** |  |  |  |
|  | a) | Report the assessment of the validity of the assumptions | 7-8 | Methods to assess the robustness of MR findings: MR Egger, MR Egger intercept test, weighted median, simple mode, weighted mode, Cochran's Q statistical test, leave-one-out analyses and *post-hoc* analyses. |
|  | b) | Report any additional statistics (e.g., assessments of heterogeneity across genetic variants, such as *I^2^*, Q statistic or E-value) | 7 | See above |
| 13 | **Sensitivity analyses and additional analyses** |  |  |  |
|  | a) | Report any sensitivity analyses to assess the robustness of the main results to violations of the assumptions | 7-8 | Methods to assess the robustness of MR findings: MR Egger, MR Egger intercept test, weighted median, simple mode, weighted mode, Cochran's Q statistical test, leave-one-out analyses and *post-hoc* analyses. |
|  | b) | Report results from other sensitivity analyses or additional analyses | 9 | By using Open Targets Genetics, we did not find other important outcomes related to our chosen genetic instruments for u-uromodulin, apart from the SNP in the UMOD gene known to be important for different renal diseases. |
|  | c) | Report any assessment of direction of causal relationship (e.g., bidirectional MR) | 9 | In the bidirectional MR analysis using upper UTIs as the exposure and u-uromodulin as the outcome, there was no suggestion of reverse causation (OR 1.02, 95% CI of 0.94-1.10, p=0.67) |
|  | d) | When relevant, report and compare with estimates from non-MR analyses | NA |  |
|  | e) | Consider additional plots to visualize results (e.g., leave-one-out analyses) |  | Leave-one-out results are presented in Supplementary table 3 |
|  | **DISCUSSION** |  |  |  |
| 14 | **Key results** | Summarize key results with reference to study objectives | 9 | In this two-sample MR study, we found that genetically predicted u-uromodulin was strongly associated with the genetically predicted risk of upper UTIs. This finding was robust across a range of sensitivity analyses. |
| 15 | **Limitations** | Discuss limitations of the study, taking into account the validity of the IV assumptions, other sources of potential bias, and imprecision. Discuss both direction and magnitude of any potential bias and any efforts to address them | 11-12 | By choosing an MR design, we greatly reduced the risk of confounding, which often affects observational studies. Furthermore, the sensitivity analyses did not indicate any bias due to pleiotropy. However, bias due to pleiotropic effects is not possible to completely rule out. In our study, this could be the case with chronic kidney disease, leading to reduced nephron mass and, hence a lower level of urinary uromodulin. The urinary uromodulin levels used in the GWAS were indexed to creatinine and the serum uromodulin levels were adjusted for eGFR. This should address most of the potential pleiotropic effects through kidney function, although there may be some bias from kidney function not captured by creatinine or eGFR. The sensitivity and subgroup analyses were affected by low statistical power. The analyses were performed on participants predominantly of European ancestry, decreasing the generalizability of our findings to the world´s other populations. A benefit of using subjects of the same ancestry group is that it reduces the risk of confounding due to population characteristics. Also, the exposure and outcome data used were from separate populations, greatly reducing the risk of confounding bias due to overlapping samples. |
| 16 | **Interpretation** |  |  |  |
|  | a) | Meaning: Give a cautious overall interpretation of results in the context of their limitations and in comparison with other studies | 11 | Our results of an association between higher genetically predicted urinary uromodulin levels and lower risk of upper UTIs support previous traditional observational studies’ findings. This strengthens the likelihood that urinary uromodulin may play a biological role in the susceptibility to upper UTIs. Further research into potential therapeutic opportunities and the possible utility of urinary uromodulin as a diagnostic marker is warranted. |
|  | b) | Mechanism: Discuss underlying biological mechanisms that could drive a potential causal relationship between the investigated exposure and the outcome, and whether the gene-environment equivalence assumption is reasonable. Use causal language carefully, clarifying that IV estimates may provide causal effects only under certain assumptions | 16 | Further, understanding of mechanisms that increase uromodulin concentrations or prevent degradation of uromodulin could potentially guide the development of new treatment options for upper UTIs. For example, a new class of small-molecular-weight compounds known as [mannosides](https://www.sciencedirect.com/topics/medicine-and-dentistry/mannoside) is being developed to treat and prevent UTIs. These agents inhibit bacterial colonization in the uroepithelium using a mechanism very similar to uromodulin |
|  | c) | Clinical relevance: Discuss whether the results have clinical or public policy relevance, and to what extent they inform effect sizes of possible interventions | 10 | The role as a biomarker would be of further importance if the level of circulating serum uromodulin was found to play a role. While there was a tendency for a protective association of increasing serum uromodulin and risk of upper UTIs, this was not significant. Earlier studies have shown that serum uromodulin protects against sepsis mortality in mice. A recently published MR study found no association between genetically determined serum uromodulin concentration and sepsis or severe pneumonia. Further studies are needed to evaluate whether measuring urinary or serum uromodulin levels can be of prognostic value when assessing patients with upper UTIs. |
| 17 | **Generalizability** | Discuss the generalizability of the study results (a) to other populations, (b) across other exposure periods/timings, and (c) across other levels of exposure | 10-11 | The analyses were performed on participants predominantly of European ancestry, decreasing the generalizability of our findings to the world´s other populations. A benefit of using subjects of the same ancestry group is that it reduces the risk of confounding due to population characteristics. Also, the exposure and outcome data used were from separate populations, greatly reducing the risk of confounding bias due to overlapping samples. |
|  | **OTHER INFORMATION** |  |  |  |
| 18 | **Funding** | Describe sources of funding and the role of funders in the present study and, if applicable, sources of funding for the databases and original study or studies on which the present study is based | 1 | KL was supported by Samarbeidsorganet Helse Midt-Norge, NTNU (Norwegian University of Science and Technology) (Trondheim, Norway). TR was funded by CTSA Grant Number UL1 TR001863 from the National Center for Advancing Translational Science (NCATS), a component of the National Institutes of Health (NIH). The contents of this manuscript are solely the responsibility of the authors and do not necessarily represent the official views of NIH. |
| 19 | **Data and data sharing** | Provide the data used to perform all analyses or report where and how the data can be accessed and reference these sources in the article. Provide the statistical code needed to reproduce the results in the article, or report whether the code is publicly accessible and if so, where |  | All data used are from publicly available resources. |
| 20 | **Conflicts of Interest** | All authors should declare all potential conflicts of interest | 12 | The authors will declare that they have no competing interests. |

This checklist is copyrighted by the Equator Network under the Creative Commons Attribution 3.0 Unported (CC BY 3.0) license.
