## Supplementary Table 2 for "UROMODULIN AND RISK OF UPPER URINARY TRACT INFECTIONS: A MENDELIAN RANDOMIZATION STUDY"

**Supplementary Table 2**: Genetic variants used as exposure for Mendelian randomization analyses

Abbreviations: rsID; reference SNP cluster ID, Chr; chromosome, EAF; effect allele frequency, EA; effect allele, OA; other allele, R^2^; squared correlation, F-statistics; fixation indices, N; number

| Exposure | Analysis | rsID | Gene | Chr | EAF | EA | OA | Beta | Standard error | P value | R^2^ | F-statistics | N |
| --- | --- | --- | --- | --- | --- | --- | --- | --- | --- | --- | --- | --- | --- |
| Urinary uromodulin | **p ≤ 5 x 10-6** | rs112907900 | C16orf82 | 16 | 0.17 | T | C | -0.26 | 0.01 | 3.86E-118 | 0.0180 | 537 | 29,315 |
|  |  | rs12815029 | TEX52 | 12 | 0.53 | T | G | 0.05 | 0.01 | 1.65E-08 | 0.0011 | 31 | 29,315 |
|  |  | rs1299147 | SNTB1 | 8 | 0.15 | A | G | -0.06 | 0.01 | 8.98E-07 | 0.0008 | 23 | 29,315 |
|  |  | rs13335818 | UMOD | 16 | 0.02 | T | C | -0.20 | 0.04 | 9.62E-07 | 0.0008 | 24 | 29,315 |
|  |  | rs1435711 | PRICKLE1 | 12 | 0.03 | T | C | 0.11 | 0.02 | 1.64E-06 | 0.0008 | 22 | 29,315 |
|  |  | rs148068503 | RNF144A | 2 | 0.82 | A | T | -0.08 | 0.02 | 1.78E-06 | 0.0008 | 24 | 29,315 |
|  |  | rs17427708 | ZBED5 | 11 | 0.02 | A | G | -0.16 | 0.03 | 1.91E-06 | 0.0008 | 22 | 29,315 |
|  |  | rs1887435 | MELK | 9 | 0.97 | A | G | 0.15 | 0.03 | 1.94E-06 | 0.0007 | 22 | 29,315 |
|  |  | rs56058634 | SLITRK5 | 13 | 0.60 | A | G | 0.05 | 0.01 | 2.01E-06 | 0.0007 | 20 | 29,315 |
|  |  | rs56359436 | FGFR2 | 10 | 0.66 | C | G | 0.04 | 0.01 | 2.07E-06 | 0.0008 | 23 | 29,315 |
|  |  | rs58315468 | WNT5A | 3 | 0.99 | T | C | -0.32 | 0.07 | 2.72E-06 | 0.0008 | 22 | 29,315 |
|  |  | rs61797906 | STXBP5L | 3 | 0.02 | A | G | 0.16 | 0.03 | 3.02E-06 | 0.0008 | 22 | 29,315 |
|  |  | rs6559619 | RPS19P6 | 9 | 0.39 | C | G | 0.05 | 0.01 | 3.49E-06 | 0.0008 | 23 | 29,315 |
|  |  | rs73375613 | TRPS1 | 8 | 0.61 | A | G | 0.04 | 0.01 | 3.64E-06 | 0.0008 | 23 | 29,315 |
|  |  | rs77056270 | CETN3 | 5 | 0.96 | A | G | -0.11 | 0.02 | 3.90E-06 | 0.0007 | 20 | 29,315 |
|  |  | rs7813392 | FGF17 | 8 | 0.05 | T | C | -0.09 | 0.02 | 3.90E-06 | 0.0007 | 21 | 29,315 |
|  |  | rs9672398 | WDR72 | 15 | 0.55 | A | G | -0.04 | 0.01 | 4.41E-06 | 0.0008 | 24 | 29,315 |
|  | **p ≤ 5 x 10-8** | rs13335818 | UMOD | 16 | 0.17 | T | C | -0.26 | 0.011 | 3.86E-118 | 0.0180 | 537 | 29,315 |
|  |  | rs9672398 | WDR72 | 15 | 0.48 | G | T | -0.05 | 0.009 | 1.65E-08 | 0.0011 | 31 | 29,315 |
| Serum uromodulin | **p ≤ 5 x 10-6** | rs117037683 | NAV3 | 12 | 0.99 | T | G | 0.32 | 0.07 | 2.19E-06 | 0.0017 | 22 | 13,556 |
|  |  | rs117608881 | THUMPD1 | 16 | 0.98 | T | C | 0.57 | 0.06 | 5.15E-21 | 0.0093 | 88 | 9,411 |
|  |  | rs11960138 | JADE2 | 5 | 0.12 | A | G | -0.10 | 0.02 | 2.19E-06 | 0.0016 | 22 | 13,956 |
|  |  | rs138462 | DDX17 | 22 | 0.96 | A | G | 0.21 | 0.04 | 3.72E-06 | 0.0022 | 21 | 9,811 |
|  |  | rs140830253 | ACSM2B | 16 | 0.98 | A | T | -0.31 | 0.06 | 2.12E-07 | 0.0029 | 27 | 9,411 |
|  |  | rs141831659 | ACSM5 | 16 | 0.98 | T | C | 0.52 | 0.06 | 2.75E-21 | 0.0065 | 90 | 13,799 |
|  |  | rs143468607 | TYRP1 | 9 | 0.97 | T | C | 0.17 | 0.04 | 3.94E-06 | 0.0015 | 21 | 13,956 |
|  |  | rs276512 | IL20RA | 6 | 0.05 | A | G | -0.14 | 0.03 | 4.54E-06 | 0.0015 | 21 | 13,713 |
|  |  | rs28510439 | UMOD | 16 | 0.11 | T | C | 0.38 | 0.02 | 3.32E-80 | 0.0251 | 359 | 13,956 |
|  |  | rs2856014 | FHIT | 3 | 0.93 | T | C | 0.12 | 0.02 | 1.11E-06 | 0.0017 | 24 | 13,956 |
|  |  | rs28562514 | ACSM5 | 16 | 0.11 | C | G | 0.12 | 0.03 | 2.44E-06 | 0.0016 | 22 | 13,956 |
|  |  | rs4500734 | UMOD | 16 | 0.98 | A | C | 0.23 | 0.04 | 6.79E-08 | 0.0021 | 29 | 13,956 |
|  |  | rs55791829 | PRKAG2 | 7 | 0.72 | C | G | -0.08 | 0.01 | 2.89E-09 | 0.0025 | 35 | 13,956 |
|  |  | rs56686587 | RNASEH2C | 11 | 0.36 | T | C | -0.07 | 0.01 | 5.11E-08 | 0.0021 | 30 | 13956 |
|  |  | rs61543910 | B4GALNT2 | 17 | 0.93 | A | T | 0.22 | 0.03 | 1.95E-15 | 0.0045 | 63 | 13,956 |
|  |  | rs62033349 | REXO5 | 16 | 0.83 | A | G | -0.09 | 0.02 | 9.29E-08 | 0.0021 | 29 | 13,713 |
|  |  | rs7224888 | B4GALNT2 | 16 | 0.9 | T | C | -0.25 | 0.02 | 1.77E-32 | 0.0100 | 140 | 13,956 |
|  |  | rs72835417 | B4GALNT2 | 17 | 0.11 | A | G | 0.20 | 0.02 | 8.36E-22 | 0.0066 | 92 | 13,956 |
|  |  | rs75044573 | UMOD | 16 | 0.04 | A | G | -0.56 | 0.04 | 1.59E-54 | 0.0173 | 242 | 13,713 |
|  |  | rs76731137 | UMOD | 16 | 0.02 | T | C | 0.26 | 0.05 | 2.08E-06 | 0.0019 | 23 | 11,584 |
|  |  | rs79303405 | ZFHX3 | 16 | 0.94 | A | G | -0.13 | 0.03 | 9.87E-07 | 0.0017 | 24 | 13,956 |
|  |  | rs8059661 | GNG13 | 16 | 0.69 | C | G | -0.11 | 0.01 | 5.13E-15 | 0.0044 | 61 | 13,956 |
|  |  | rs9928757 | GP2 | 16 | 0.19 | C | G | -0.69 | 0.02 | 1.00E-200 | 0.1312 | 2107 | 13,956 |
|  |  | rs9983032 | NRIP1 | 21 | 0.86 | C | G | -0.08 | 0.02 | 4.18E-06 | 0.0015 | 21 | 13,956 |
