## Supplementary Table 3 for "UROMODULIN AND RISK OF UPPER URINARY TRACT INFECTIONS: A MENDELIAN RANDOMIZATION STUDY"

**Supplementary Table 3:** Leave-one-SNP-out analysis in the Mendelian randomization analysis of urinary uromodulin.

Abbreviations: SNP; single nucleotide polymorphism, OR; odds ratio

| SNP left out | OR (95% CI)^a^ | P value |
| --- | --- | --- |
| rs112907900 | 0.77 (0.65-0.92) | 0.004 |
| rs12815029 | 0.80 (0.67-0.96) | 0.014 |
| rs1299147 | 0.81 (0.68-0.97) | 0.020 |
| rs13335818 | 0.78 (0.60-1.02) | 0.069 |
| rs1435711 | 0.79 (0.66-0.94) | 0.009 |
| rs148068503 | 0.80 (0.67-0.96) | 0.016 |
| rs17427708 | 0.78 (0.65-0.93) | 0.006 |
| rs1887435 | 0.80 (0.67-0.96) | 0.017 |
| rs56058634 | 0.79 (0.66-0.94) | 0.009 |
| rs56359436 | 0.79 (0.66-0.94) | 0.010 |
| rs58315468 | 0.79 (0.66-0.95) | 0.010 |
| rs61797906 | 0.80 (0.67-0.96) | 0.014 |
| rs6559619 | 0.80 (0.67-0.96) | 0.015 |
| rs73375613 | 0.81 (0.67-0.96) | 0.016 |
| rs77056270 | 0.80 (0.67-0.96) | 0.016 |
| rs7813392 | 0.80 (0.67-0.96) | 0.014 |
| rs9672398 | 0.81(0.68-0.97) | 0.019 |

^a^The odds ratios (OR) correspond to a one standard deviation increase in the concentration of urinary uromodulin.
